# Mathematical modeling of COVID-19 transmission and mitigation strategies in the population of Ontario, Canada

**DOI:** 10.1101/2020.03.24.20042705

**Authors:** Ashleigh R. Tuite, David N. Fisman, Amy L. Greer

## Abstract

**Background:** We evaluated how non-pharmaceutical interventions could be used to control the COVID-19 pandemic and reduce the burden on the healthcare system.

**Methods:** Using an age-structured compartmental model of COVID-19 transmission in the population of Ontario, Canada, we compared a base case with limited testing, isolation, and quarantine to scenarios with: enhanced case finding; restrictive social distancing measures; or a combination of enhanced case finding and less restrictive social distancing. Interventions were either implemented for fixed durations or dynamically cycled on and off, based on projected ICU bed occupancy. We present median and credible intervals (CrI) from 100 replicates per scenario using a two-year time horizon.

**Results:** We estimated that 56% (95% CrI: 42-63%) of the Ontario population would be infected over the course of the epidemic in the base case. At the epidemic peak, we projected 107,000 (95% CrI: 60,760-149,000) cases in hospital and 55,500 (95% CrI: 32,700-75,200) cases in ICU. For fixed duration scenarios, all interventions were projected to delay and reduce the height of the epidemic peak relative to the base case, with restrictive social distancing estimated to have the greatest effect. Longer duration interventions were more effective. Dynamic interventions were projected to reduce the proportion of the population infected at the end of the two-year period. Dynamic social distancing interventions could reduce the median number of cases in ICU below current estimates of Ontario’s ICU capacity.

**Interpretation:** Without significant social distancing or a combination of moderate social distancing with enhanced case finding, we project that ICU resources would be overwhelmed. Dynamic social distancing could maintain health system capacity and also allow periodic psychological and economic respite for populations.

## Background

The COVID-19 pandemic represents a global public health emergency unparalleled in recent time. In the two months since the initial World Health Organization report describing the COVID-19 outbreak concentrated in Wuhan City, China (1), the number of confirmed cases has risen sharply from 282 to more than 330,0000, with 14,510 reported deaths across all regions of the globe (2). The first imported case of COVID-19 in Ontario, Canada was reported on January 25, 2020 and community transmission was first documented on March 1, 2020 in British Columbia, Canada (3).

This pathogen represents a significant challenge for public health, pandemic planning, and healthcare systems. The SARS-CoV-2 virus is highly transmissible (4-7). It causes moderate to severe clinical outcomes in approximately 20% of all recognized infected individuals (5, 8, 9). In the absence of a vaccine, public health responses have focused on the use of non-pharmaceutical interventions (NPIs) (10). These NPIs include: (1) “case-based” measures such as testing, contact tracing, isolation, and quarantine, and (2) “non-case-based” measures such as reducing the probability of transmission given an effective contact (e.g. hand hygiene, and cough etiquette), and social distancing measures to reduce the contact rate in the population. Social distancing minimizes opportunities for person-to-person transmission of the virus to occur. These social distancing measures include some combination of school closure, teleworking, cancellation of group activities and events, and a general overall reduction in community contacts. While they are expected to be effective in reducing transmission of SARS-CoV-2, they are also associated with substantial economic costs and social disruption.

Epidemiological models can contribute important insight for public health decision-makers by allowing for the examination of a variety of “what-if” scenarios. The Canadian Pandemic Influenza Plan (CPIP) for the Health Sector (the backbone of which informs COVID-19 pandemic preparedness and response) identifies two main objectives for responding to a pandemic: (1) to minimize serious morbidity and mortality, and (2) to minimize societal disruption (11). The overarching goal of pandemic response is to find a combination of NPIs that would minimize the number of cases requiring in-patient medical care (e.g. hospitalization and /or ICU admission), and deaths, while also minimizing the level of societal disruption.

Societal disruption measures could include outcomes such as the overall duration of time that the intervention needs to be to achieve the associated reductions in morbidity and mortality. A challenge for pandemic response using NPIs is that, in a fully susceptible population, although NPIs may slow disease transmission while they are in place, once the intervention is lifted (or compliance with the intervention becomes low), the transmission of the pathogen rebounds rapidly (10, 12). In the case of COVID-19, it may not be possible to minimize morbidity and mortality, and societal and economic disruption at the same time.

Given these considerations, we used a transmission dynamic model of COVID-19 to explore the potential impact of case-based, and non-case-based NPIs in the population of Ontario, Canada. Our analysis focusses on identifying strategies that keep the number of projected severe cases (hospitalizations, and ICU admissions) within a range that would not overwhelm the Ontario healthcare system, while also considering the amount of time these interventions would be in place.

## Methods

### Model overview

We developed an age-structured compartmental model that describes COVID-19 transmission in the province of Ontario, Canada. We used a modified ‘Susceptible-Exposed-Infectious-Recovered’ framework that incorporated additional compartments to account for public health interventions, different severities of clinical symptoms, and hospitalization risk. An overview of the model compartments and movements between them is provided in **Figure 1** and model equations and additional details are provided in the **Technical Appendix**. The model was run for a period of two years and we assumed that recovered individuals remain immune from re-infection for the duration of the epidemic. Individuals remained infectious until they recovered or were hospitalized; we did not model transmission within healthcare settings. For simplicity, we assumed that all deaths occurred in cases requiring intensive care. We included cases in hospital and requiring intensive care to estimate health care requirements over the course of the epidemic. The model was constructed in R (13).

**Figure 1.**
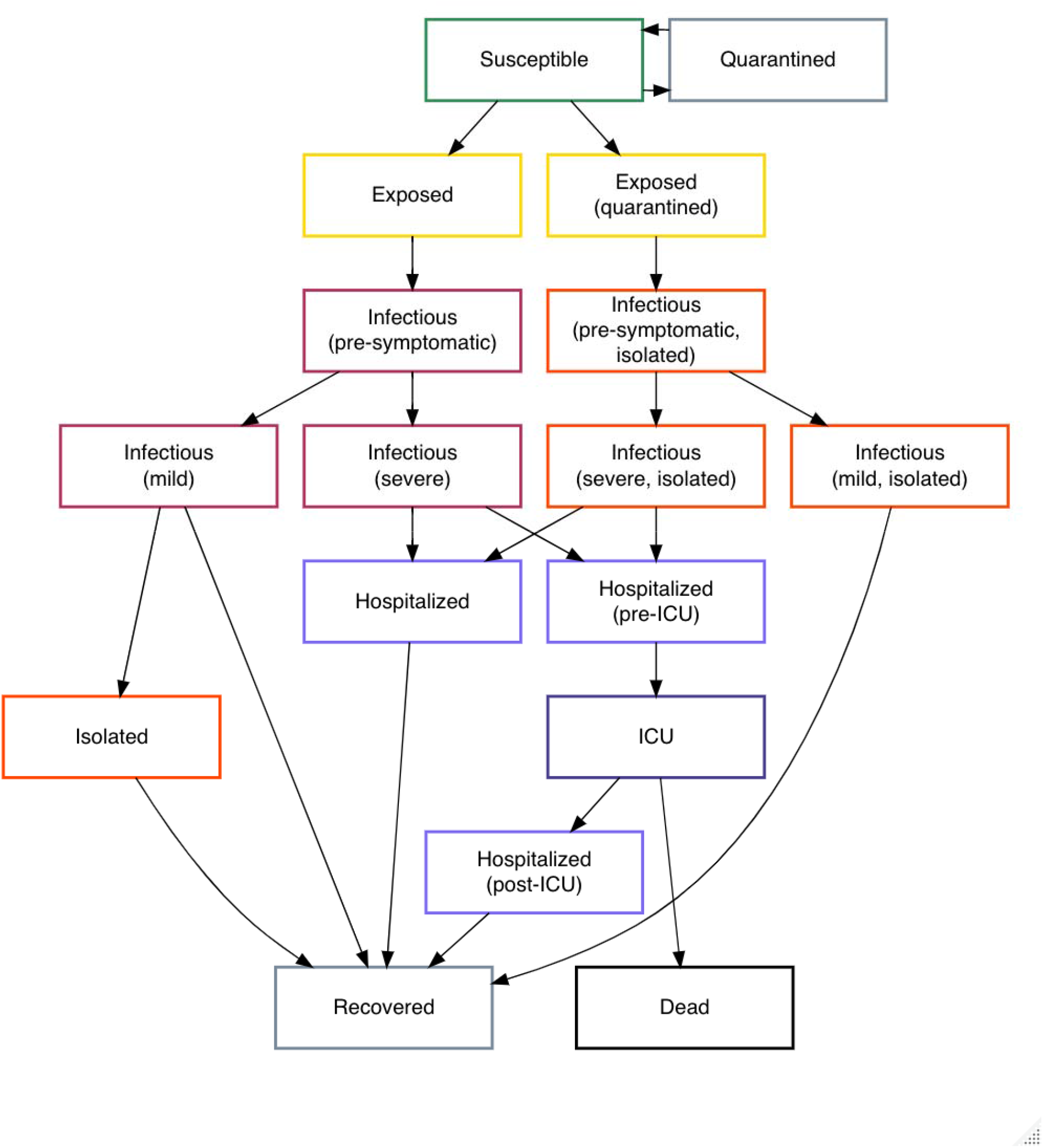
COVID-19 transmission model structure. Exposed cases can be either quarantined or not; quarantined cases would represent those who were identified via contact tracing. Hospitalized cases are assumed to be no longer infectious to others due to recognition of infection and are included in the model to estimate healthcare requirements. The model is stratified by age group and presence /absence of comorbidities.

### Model parameters

The model was stratified by 5-year age groups using 2019 population estimates (14). Contacts within and between age groups were based on the POLYMOD study (15), using contact data specific for the United Kingdom. The model was further stratified by health status to account for differential vulnerability to severe infection among those with underlying health conditions. We obtained comorbidity estimates by age from the Canadian Community Health Survey (16) for Ontario and included the following conditions: hypertension, heart disease, asthma, stroke, diabetes, and cancer. For younger age groups (<12 years) we used estimates from Moran et al. (17).

Parameters describing the natural history and clinical course of infection were derived from published studies (**Table 1**, full details in **Technical Appendix**). To capture variability in transmission, specifically the observation that the basic reproductive number (R0) is over-dispersed, with some cases transmitting to many others (superspreader events), while many other cases transmit much less, we have added volatility to the transmission term (18). The model was initiated with 750 prevalent cases (based on 150 reported cases in Ontario on March 19, 2020 and an assumed reporting rate of 20%), that were randomly distributed across the infectious compartments.

**Table 1.**
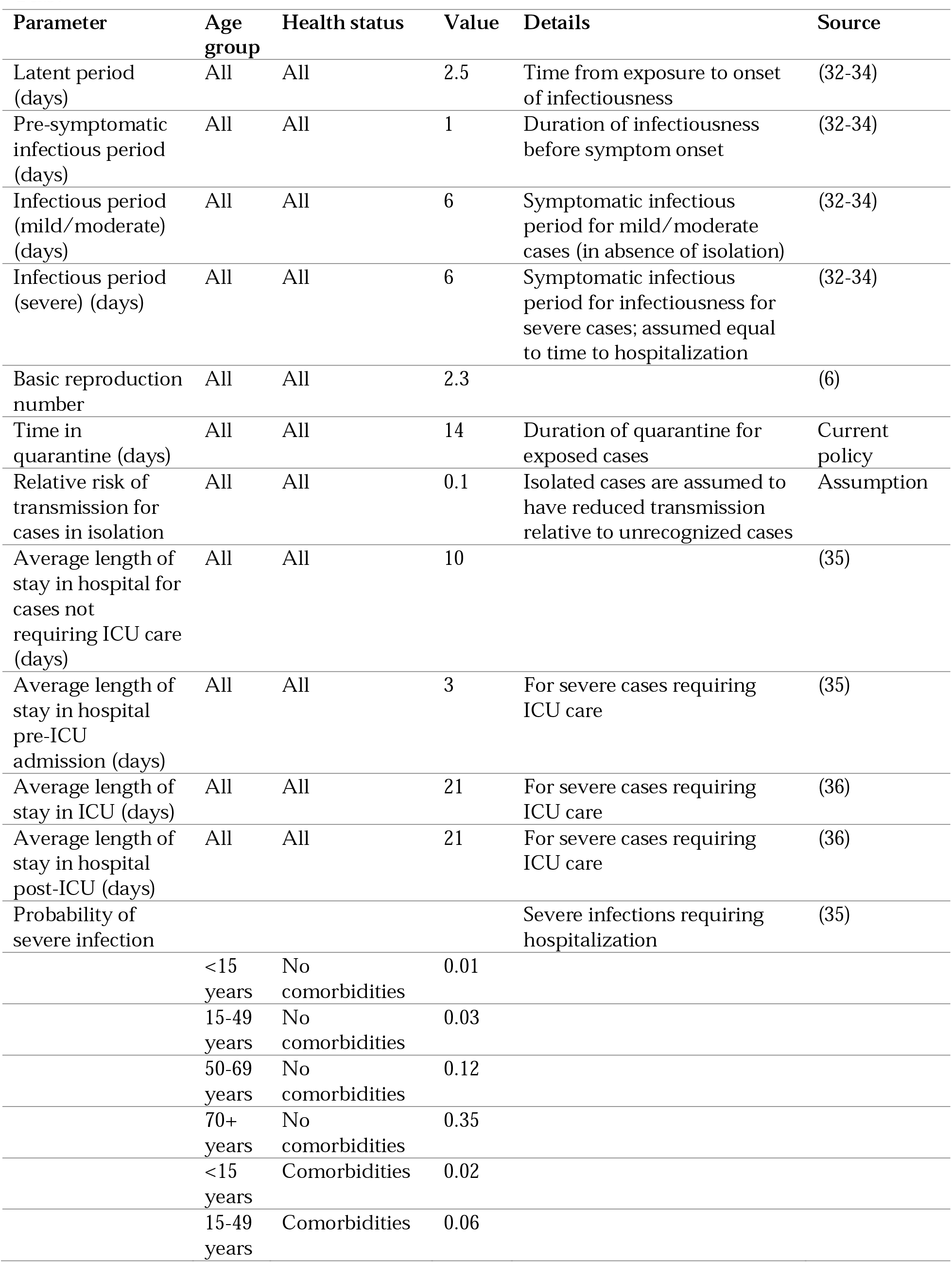
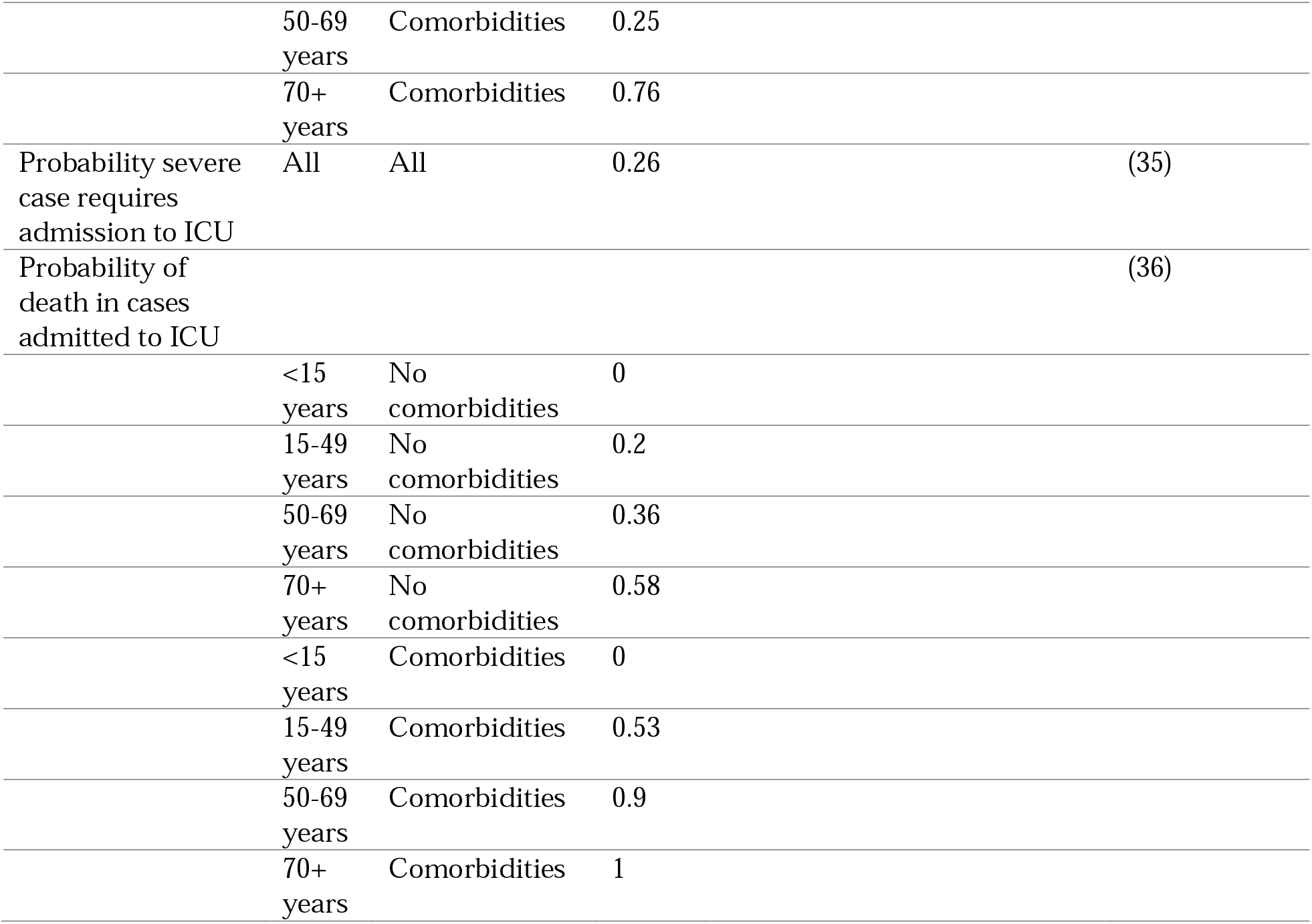
Model parameters.

### Interventions

Testing was assumed to move individuals with non-severe symptoms from the infectious to isolated compartments. Isolated cases were assumed to have reduced transmission compared to non-isolated cases. Social distancing measures were assumed to reduce the number of contacts per day across the entire population. Details of parameters that were varied under different interventions are included in **Table 2**. For the base case, we assumed that there was a degree of testing and isolation occurring and that a proportion of exposed cases were quarantined. We then added in additional control measures: (i) enhanced testing and contact tracing; (ii) restrictive social distancing measures; and (iii) a combination of enhanced testing and contract tracing, along with less restrictive social distancing than in (ii). We considered two approaches to implementing interventions: (i) fixed durations and (ii) a dynamic approach with interventions turned on and off based on the number of cases requiring ICU care in the population. We focused on ICU capacity, since this is expected to be most limited resource during the COVID-19 epidemic. Prior to emergence of COVID-19, Ontario had approximately 1300 ICU beds with associated ventilators; but 90% were occupied by individuals with non-COVID-19 illness; in mid-March 2020, the Ontario government made 300 additional ventilator-associated ICU beds available (for a total of 430 unoccupied beds). As such, we used 200 COVID-19 cases in the ICU (across all of Ontario) as a threshold for turning the intervention on. This value was based on ~50% saturation of available beds combined with the recognition that there is a lag between cases acquiring infection and requiring intensive care, such that one would expect ICU needs to grow rapidly once initial COVID-19 cases present for care.

**Table 2.** Model scenarios details.

| Parameter | Age group | Scenario |  |  |  |
| --- | --- | --- | --- | --- | --- |
|  |  | Base case | Enhanced case detection | Social distancing | Enhanced detection with limited social distancing |
| Non-quarantined cases tested and isolated (%) |  |  |  |  |  |
|  | <15 years | 10 | 40 | 10 | 40 |
|  | 15 – 49 years | 40 | 60 | 40 | 60 |
|  | ≥50 years | 70 | 80 | 70 | 80 |
| Exposed cases in quarantine before infectious (%) | All | 10 | 30 | 10 | 30 |
| Reduction in contacts with social distancing (%) | All | 0 | 0 | 60 | 25 |

### Outputs

Key model outputs included final epidemic attack rates (% of population infected at the end of the 2-year period), prevalence of hospitalizations and ICU use, and deaths. For comparison we show the maximum and current ICU capacity per 10,000 population relative to model projections. For the dynamic intervention scenarios, we also calculated the amount of time over the 2-year model period during which the intervention was implemented, as a measure of intervention intensity. We present model outputs as medians and credible intervals (CrI) from 100 model replicates per intervention.

## Results

### Base case

In the model base case, with limited testing, isolation and quarantine, we estimated that 56% (95% CrI: 42-63%) of the Ontario population would be infected over the course of the epidemic. This would include cases of all severities. Attack rates were projected to be highest in those aged 5-14 years (77%, 95% CrI: 63-83%) and 15-49 years (63%, 95%CrI: 48-71). Lower attack rates were projected in individuals aged less than 5 years (50%, 95% CrI: 37-58%) and adults aged 50-69 years (47%, 95% CrI: 34-55) and greater than 70 years (30%, 95% CrI: 21-36). An example of the outbreak trajectory across model simulations is presented in **Figure 2**. At the peak of the epidemic, in the absence of any resource constraints to provide care (i.e., assuming all cases requiring medical care receive it), we projected 107,000 (95% CrI: 60,760-149,000) cases in hospital and 55,500 (95% CrI: 32,700-75,200) cases in ICU. The high prevalence of cases in ICU reflects the mean length of ICU stay associated with COVID-19 infection in other countries.

**Figure 2.**
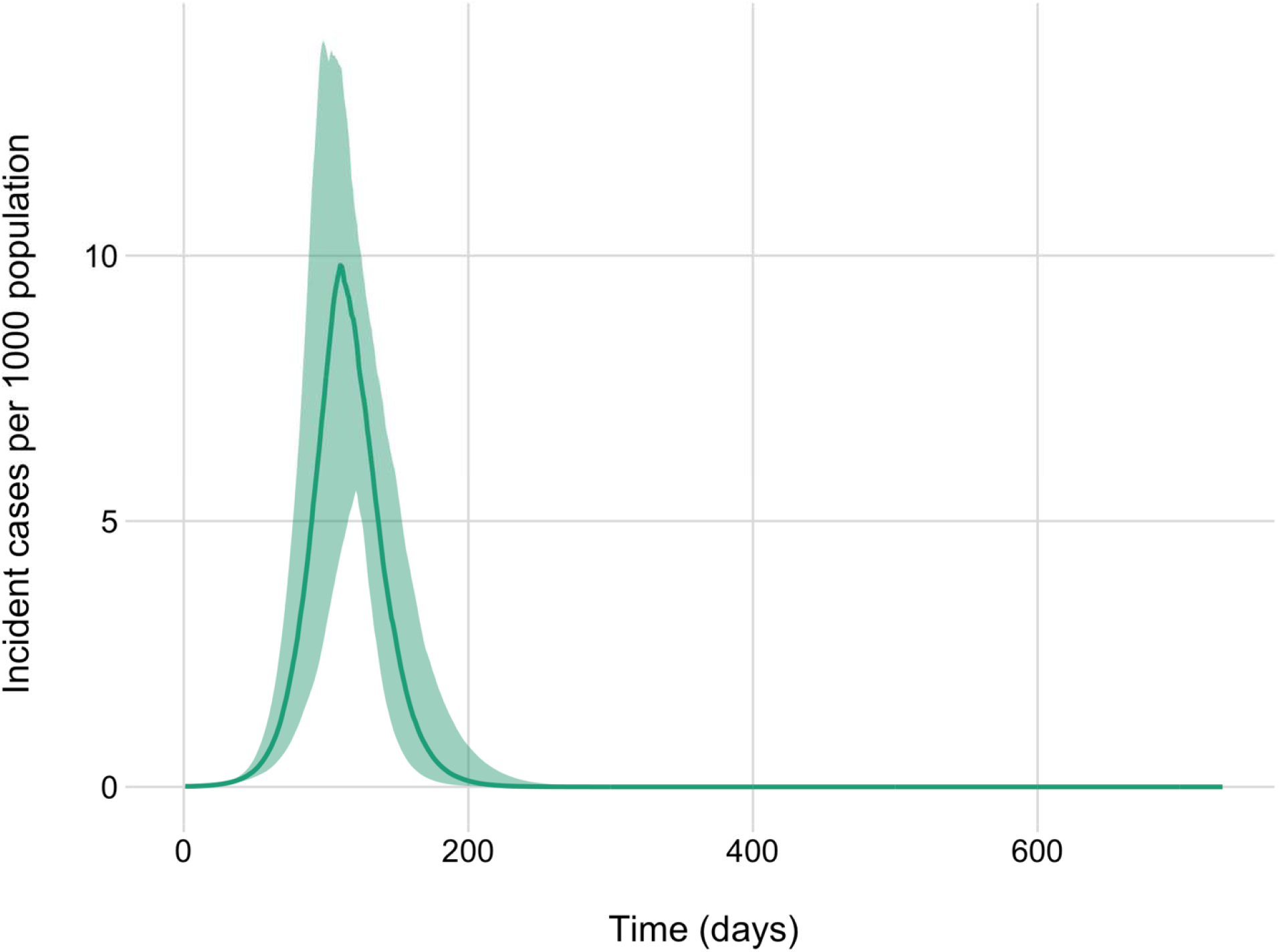
Projected COVID-19 epidemic trajectory for the base case model with minimal intervention. Daily incident cases per 1000 population are presented. The line represents the median value of 100 model simulations and the shaded area indicates the 95% credible interval.

### Fixed duration interventions

All of the interventions considered were projected to delay the epidemic peak and reduce the number of cases requiring ICU care at the peak (**Figure 3**). The effectiveness of the interventions scaled with intervention duration. For all interventions, when the intervention duration was 6 months or less, there was no appreciable difference on final attack rate. With 12 and 18 months of heightened response measures, the proportion of the population infected at the end of the 2-year period was reduced and, in some simulations, the prevalence of cases requiring intensive care fell below Ontario’s current capacity for all or part of the time period. The largest effect was observed for the restrictive social distancing intervention. The combination intervention, with enhanced case detection and less aggressive social distancing was projected to substantially reduce attack rates when implemented for 18 months, while enhanced case detection in the absence of social distancing measures had a more modest effect, on average. There was substantial variability in model projections, due to model stochasticity.

**Figure 3.**
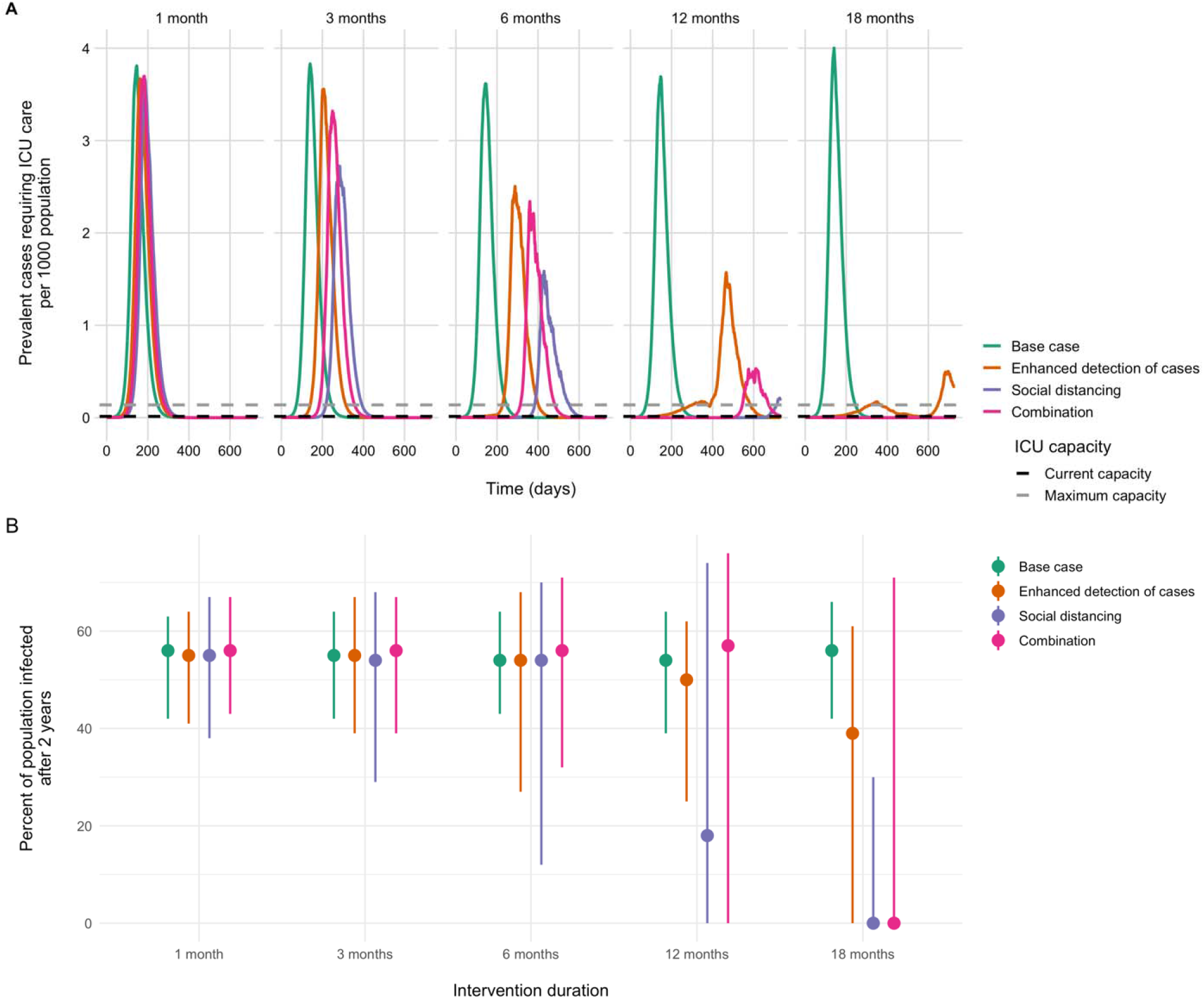
Projected ICU bed requirements and attack rates for fixed duration interventions. (A) Prevalent cases requiring intensive care are shown for intervention durations of 1, 3, 6, 12, and 18 months. Maximum and current ICU capacity in Ontario are indicated by the dashed horizontal lines. Median values are presented. (B) Model-projected percent of the population infected over the 2-year time period. Attack rates include all infections, regardless of severity. Note that the slight variability in epidemic size for the no additional intervention base case reflects model stochasticity across simulations.

### Dynamic interventions

We also explored dynamic interventions that were turned on and off in response to the current state of the epidemic. Dynamic interventions were projected to be effective for reducing the proportion of the population infected at the end of the two-year period, with potentially shorter durations of social distancing than the fixed duration approach (**Figure 4)**. For example, when implemented dynamically, 13 months of social distancing, cycled on and off, reduced the mean overall attack rate to 2%. For the social distancing alone and combination intervention scenarios, we observed atypical epidemic curves, with the number of cases increasing and decreasing repeatedly over time. In these scenarios, the median number of cases in ICU was reduced below current estimates of Ontario’s ICU capacity.

**Figure 4.**
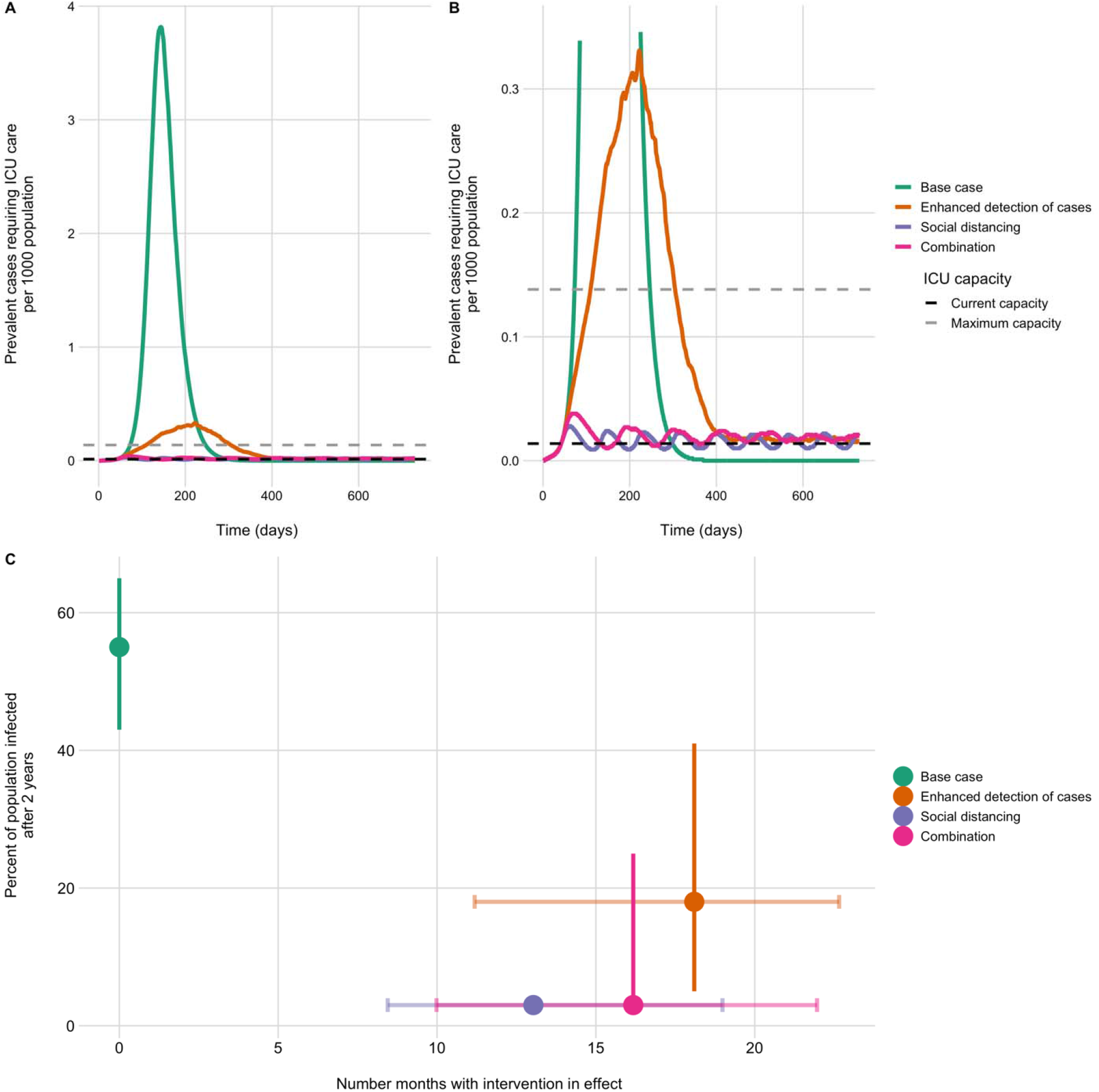
Projected ICU bed requirements and attack rates for dynamic interventions. (A) Prevalent cases requiring intensive care are shown for the base case and three intervention scenarios. Interventions are turned on and off (returning to base case parameter values), depending on the number of COVID-19 cases in the ICU. Maximum and current ICU capacity in Ontario are indicated by the dashed horizontal lines. Median values are presented. (B) Zoomed view of prevalent ICU cases to show the dynamics for the enhanced social distancing and combination scenarios. (C) Model-projected estimates of percent of the population infected over the 2-year time period. Attack rates include all incident infections, regardless of severity. Median and 95% CrI are shown. The amount of time the dynamic interventions are in place is shown on the x-axis. Points indicate the median duration and lines the 95% CrI for each scenario.

## Interpretation

COVID-19 poses an extraordinary challenge to societies. While severe illness, particularly in older individuals, is frequent enough to overwhelm a society’s ICU capacity (19), mild unrecognized illness (particularly in younger individuals) contributes to spread (20), and epidemics may only be recognized when superspreader events occur (21), often in vulnerable settings like health care facilities (22). In contrast to SARS (23), the high frequency of mild cases means that strategies which focus on case identification and isolation alone are likely to fail (22). As such, population-level interventions, with their attendant economic costs, have been used to prevent health systems from collapsing (24). While events in China, Singapore, Hong Kong and elsewhere have demonstrated that COVID-19 epidemics can be contained (24-27), the seeding of epidemics in countries around the globe, many with weak health systems (28), means that reintroduction of COVID-19 will continue to occur for some time. As successful containment efforts maintain a large number of susceptible individuals in populations, vulnerability to repeated epidemics is likely to persist until a COVID-19 vaccine is developed and manufactured at scale; or until large fractions of the population are infected and either die or develop immunity (29).

Control strategies for COVID-19 thus need to balance competing risks: the risks of mortality and health system collapse, on the one hand, against economic risks and attendant hardships (and health consequences) on the other. In this work, we evaluated plausible strategies for attenuating the COVID-19 epidemic in Ontario, Canada. We focussed on ICU resources for two reasons: first, because this component of most health systems represents a scarce resource prone to being saturated; and secondly because such saturation results in abrupt surges in case-fatality, as individuals with acute respiratory distress syndrome will die quickly without the capacity for mechanical ventilation. In broad terms, we find that prolonged social distancing is the preferred strategy for maintaining ICU resources, but an extreme fixed duration of social distancing is required to prevent the epidemic from overwhelming ICU capacity. That said, social distancing, even without reducing overall outbreak size, has the added benefit of delaying the epidemic peak, which gains time that can be used to build health system capacity and identify therapies and vaccines.

In contrast to fixed-duration social distancing, we find that dynamic social distancing, with interventions turned on and off as needed, based on ICU capacity crossing a given threshold, represents a more effective, and likely more palatable, control strategy. Social distancing can be relaxed, but this inevitably results in resurgent disease in the population, requiring reinstatement. Nonetheless, dynamic social distancing is projected to maintain ICU capacity, and dramatically reduces overall attack rates, while at the same time requiring less total social distancing time than would be required by a fixed duration strategy of comparable effectiveness. Furthermore, dynamic social distancing has the potential to allow populations, and the economy, to “come up for air” at intervals, which may make this strategy more sustainable. We also found that a combination approach, with less restrictive social distancing along with enhanced case isolation and quarantine, could have a similar effect in the dynamic scenario as more restrictive social distancing alone. It is plausible that, as testing capacity increases, a combination approach that is less reliant on social distancing, will strike the right balance between disease control and societal disruption (30).

Like any model, ours has limitations. At the time of writing, limitations in testing capacity in Ontario, and lack of information on ICU occupancy by COVID-19 patients, makes it challenging to know where exactly on the epidemic curve we currently find ourselves. Any model involves trade-offs between simplicity and realism, and in the current work we have not attempted to model social distancing measures in a highly realistic way, but rather generically as reductions in contact frequency. Our understanding of the natural history of SARS-CoV-2 infection continues to evolve and the precise role of pre-symptomatic and subclinical transmission is uncertain. Social distancing becomes a more important control measure in the face of incomplete case ascertainment due to asymptomatic or mildly symptomatic cases. The model does not include seasonality; it is possible that transmission will attenuate in the summer (31), resulting in a decline in cases that would be expected to resurge with the return of colder weather. All of these factors mean that the quantitative findings are subject to uncertainty.

Nonetheless, the qualitative insights around the role of social distancing, the relatively long intervention durations required to bend the epidemic curve, and the potential use of cyclic interventions can be used to policy-makers and decision makers, along with emerging empirical evidence from other countries, to consider the best approaches for epidemic control over the coming months. Lastly, we have not modeled the fact that that abrupt surges in death resulting from full ICUs would, result in lower demands for ICU beds. Our goal here is to inform policy so that such outcomes are avoided to the extent possible.

In summary, we have modelled plausible contours of the COVID-19 epidemic in Ontario, Canada, with a focus on maintenance of ICU resources. In the absence of significant social distancing or a combination of moderate social distancing with enhanced case detection and isolation, we project that ICU resources would be quickly overwhelmed, a conclusion consistent with that in other modeling work (12), as well as current events in Italy and Spain. On a more positive note, we project that dynamic social distancing, that reacts to changes in ICU occupancy, could maintain health system capacity and also allow periodic psychological and economic respite for populations.

## Data Availability

Data are available from the corresponding author on request.

## Acknowledgements

The authors wish to thank Gabrielle Brankston, Shannon French, Tanya Rossi, and Matthew Van Camp from the Department of Population Medicine, University of Guelph for helping to compile data on population demographics and chronic conditions. The authors also acknowledge the feedback of colleagues from local public units across the province of Ontario.

